# Abortion rights for refugees around the world: A scoping review and quantitative analysis of abortion laws and forced displacement in 2022

**DOI:** 10.1101/2025.01.16.25320653

**Authors:** Blake Erhardt-Ohren, Ndola Prata

## Abstract

**Background:** The refugee population increases around the world each year, and 25% of these individuals are capable of pregnancy. We sought to understand the potential impact to refugees’ lives, with regard to their ability to access abortion services, as they crossed international borders.

**Methods:** We analyzed global refugee movement in 2022 and country-level abortion law data to identify patterns of forced migration. We conducted a scoping review to identify emerging topics and research gaps related to refugee fertility intentions and access to reproductive health services.

**Results:** 6,860,398 female refugees aged 12-59 years moved between countries in 2022. 76% of these refugees went to host countries without abortion on request. 16% of these refugees sought asylum in countries with more restrictive abortion laws than their country of origin, 44% sought asylum in countries with similarly restrictive laws, and 40% sought asylum in countries with less restrictive laws. We identified fifty-two items in our scoping review with the following themes: unintended fertility; intended fertility; need to space, postpone and/or limit fertility; changing fertility intentions; and access to reproductive health services. These items revealed mixed fertility intentions among newly resettled adult refugees and clear evidence that adolescent refugees want to control their fertility despite significant barriers.

**Discussion:** The results reveal the need for renewed commitment to ensuring that refugees capable of pregnancy have access to the full range of sexual and reproductive health services, so they can exercise their human right to decide if, when, and how many children to have.

**Plain language summary:** The refugee population increases each year and approximately 25% of refugees are capable of pregnancy. The purpose of this study is to understand how refugee status affects refugees’ ability to access abortion. We analyzed refugee movement in 2022 and countries’ abortion law data to identify patterns of forced migration and associated changes in law. We conducted a scoping review to identify emerging or evolving topics and research gaps related to refugee fertility intentions and access to reproductive health services. 6,860,398 female refugees aged 12-59 years moved from 159 origin countries to 111 host countries in 2022. Around 76% of these refugees went to host countries that did not provide abortion on request: 16% of these refugees sought asylum in countries with more restrictive abortion laws than their country of origin, 44% sought asylum in countries with similarly restrictive abortion laws, and 40% sought asylum in countries with less restrictive abortion laws. Fifty-two items in our scoping review revealed mixed fertility intentions among newly resettled adult refugees and clear evidence that adolescent refugees want to control their fertility, despite significant barriers. The study results point to the need for a renewed commitment to ensuring that refugees capable of pregnancy have access to the full range of sexual and reproductive health services, so they can exercise their human right to decide if, when, and how many children to have.

## Background

The number of new refugees grows around the world each year and shows no sign of slowing.^1^ When the United Nations High Commissioner for Refugees (UNHCR) began reporting annual numbers of refugees in 1951, they totaled 2,116,011; at the end of 2022, the number had grown to 29,413,033, an increase of about 1,290%.^2^ Between 2017 and 2022 alone, the number grew by almost 10 million individuals. These numbers are likely underestimates, not overestimates.^3^ Not only does the number of forcibly displaced individuals continue to grow, but the causes of displacement are also increasing. The factors accelerating forced displacement globally are well-known. Pre-existing drivers, such as those related to the environment (i.e. drought), society (i.e. lack of opportunity for community members), politics and policy (i.e. corruption), and economy (i.e. poverty), are triggered by immediate events, such as armed attacks or natural disasters, that then force individuals to flee their homes.^4^ The most common culprit, however, is political, and may both trigger and protract crises through creation of structural weaknesses and misappropriation of aid.^4^

One in four of the almost 30 million refugees at the end of 2022 is an individual who can become pregnant.^1^ These refugees may have a higher unmet need for high-quality, accessible safe abortion and post-abortion services than the general population due to the constraints of service provision in complex humanitarian emergencies. Health facilities in these settings often experience stockouts of contraceptive methods or may not have the providers or equipment necessary to provide high-quality reproductive health services.^5^ This reduced access to commodities and services may increase the number of unintended pregnancies in these populations.^6^ Additionally, refugees are more likely to experience rape, early or forced marriage, intimate partner violence, child sexual abuse, prostitution, sex trafficking, or other forms of sexual violence and exploitation than communities or individuals in relatively stable settings.^7^ In complex humanitarian emergencies, the reported prevalence of sexual-based violence is 21.4%^8^ compared to about 10% globally.^9^ Decreased access to contraception, coupled with increased sexual violence, puts refugees at a higher risk of unintended pregnancy.

Once refugees become pregnant, they may face barriers to safe abortion care if they wish to end their pregnancy. A scoping review of access to safe abortion care in humanitarian crises found that barriers include those related to the legal environment, context, stigma, economic factors, and service delivery.^10^ Potential reasons that refugees cannot access or do not use comprehensive abortion care include barriers at the policy, community, health facility, interpersonal, or individual level.^11–14^

Many countries have signaled their protection of the right to safe and legal abortion services and post-abortion care for complications by signing the United Nations’ international treaties that interpret the Universal Declaration of Human Rights (UDHR), such as the Convention on the Elimination of All Forms of Discrimination against Women (CEDAW), the Convention against Torture and Other Cruel, Inhuman or Degrading Treatment or Punishment (CAT), and many others.^15^ Many of these treaties target protections for vulnerable individuals, and specifically people who have been forcibly displaced. The safeguards conferred in the UDHR and interpreted in international treaties have been used to hold states accountable for violations of human rights related to abortion, such as in recent cases in Croatia (2022),^16^ Ireland (2016-2017),^17^ and Chile (2015).^18^

Beyond treaties, there are other ways that civil societies and academic institutions have worked to improve the reproductive health of forcibly displaced persons. Several civil society organizations and academic institutions have recognized the right to reproductive health of forcibly displaced persons and developed agendas to improve their reproductive health and access to quality services, such as through the Inter-Agency Working Group on Reproductive Health in Crises and the RAISE Initiative.^19,20^ These and other actors in the humanitarian space have called for a justice lens to push for a rights-based approach to service implementation.^21^ To improve visibility of forcibly displaced persons, in March 2024, the UN Statistical Commission unanimously endorsed a recommendation to improve the inclusion of refugees and internally displaced persons in national data systems.^22^ Additional research has called out the need for evaluation of reproductive health programs targeting forcibly displaced persons,^23^ especially in fulfillment of the Sustainable Development Goals,^24^ and documented progress since the 1994 International Conference for Population and Development and ongoing needs for humanitarian response.^25^

The purpose of this study is to provide evidence of the potential impact to refugees’ lives with regard to their ability to access safe and legal abortion, as they crossed borders to seek protection. We believe that it is important to understand, at a high level, what percent of the population of refugees capable of pregnancy lost their right to abortion by due to forced migration. To do this, we use data from 2022 to estimate how many refugees may have become more vulnerable by moving to a country with more restrictive abortion laws relative to their country of origin. To contextualize the quantitative findings, we review current evidence on refugee fertility intention and access to comprehensive reproductive health services to better understand the impact that these laws have on refugees’ health and well-being. The overall goal of this study is to provide evidence of the need to pay particular focus to abortion rights and access for refugee women and girls.

## Materials and methods

### Definitions

For the purpose of this study, we define refugees according to the UNHCR’s Data Finder, which includes both refugees and “people in refugee-like situation”.^2^ Refugees are individuals who are protected under the 1951 Convention on the Status of Refugees (and its 1967 Protocol), the 1969 Organization of African Unity Convention Governing the Specific Aspects of Refugee Problems in Africa, the 1984 Cartagena Declaration on Refugees, the UNHCR Statute, and individuals that are granted complementary forms of protection or under temporary protection.2 They are defined by the United Nations as [individuals, who] “owing to well-founded fear of being persecuted for reasons of race, religion, nationality, membership of a particular social group or political opinion, is outside the country of his nationality and is unable or, owing to such fear, is unwilling to avail himself of the protection of that country; or who, not having a nationality and being outside the country of his former habitual residence, is unable or, owing to such fear, is unwilling to return to it.”^26^ People in refugee-like situation are individuals residing outside of their country or territory of origin who face similar circumstances as refugees, but who have not been given refugee status for practical or other reasons. In this manuscript, we refer to both refugees and people in refugee-like situation as refugees. We use “origin country” to mean the country from which a refugee fled and “host country” to mean the country to which a refugee fled and where they currently reside. We preserved the country names in the UNHCR Refugee Data Finder query and use those country names throughout this manuscript.^2^

#### Quantitative analysis

##### Refugee data

We retrieved data from the UNHCR Refugee Data Finder^2^ on 03 August 2023 for the calendar year 2022. The data query included aggregate data with origin country, host country, sex, and age group (0-4, 5-11, 12-59, 60+ years, other) for refugees only. Due to data constraints, we were unable to disaggregate data for women of reproductive age (WRA), 15-49 years^27^ and unable to extrapolate the proportion of refugees who may be WRA. We included data for females 12-59 years old and excluded all male data and female data for the age group labeled “other”. We further excluded refugees with “Unknown” or “Stateless” origin country due to our inability to assign abortion laws for these locations.

##### Abortion law data

We matched the columns for origin country and host country in the UNHCR Refugee Data Finder query to the countries included in the Center for Reproductive Rights (CRR)’s The World’s Abortion Laws map.^28^ At this point, we also removed “China, Hong Kong SAR” and “Western Sahara” as origin countries and “China, Hong Kong SAR” and “Western Sahara” as host countries, due to their missingness in the CRR’s The World’s Abortion Laws map. We matched each country in the UNHCR data query to its corresponding abortion law category based on the CRR designation, which included “prohibited altogether”, “to save the pregnant person’s life”, “to preserve health”, “socioeconomic grounds”, “on request”, and “varies by state”.^28^ We reviewed the Council on Foreign Relations’ summary of changes to abortion law between 2000 and 2022,^29^ then reviewed the World Health Organization’s Global Abortion Policies Database for each of the 38 listed countries to confirm the year the law changed and when the law entered into force.^30^

##### Further exclusion criteria

Within the merged data set, we excluded countries where new abortion laws entered into force during the 2022 calendar year, due to our inability to disaggregate between individuals that became refugees before and after law changes in their host country.

##### Data analysis

We assigned each abortion law category a number based on severity, with 5 being the most severe: “prohibited altogether” = 5, “to save the pregnant person’s life” = 4, “to preserve health” = 3, “socioeconomic grounds” = 2, and “on request” = 1. We excluded countries that were categorized by the CRR as “varies by state”. Finally, we determined whether individuals traveled from countries with more restrictive laws to countries with less restrictive laws, from countries with less restrictive laws to countries with more restrictive laws, or if the abortion law category was the same in the individuals’ origin country and host country. We analyzed the data using the dplyr^31^ and ggplot2^32^ packages in R programming^33^ to produce descriptive statistics.

#### Scoping review

We conducted a scoping review to complement the quantitative data analysis. The scoping review is not limited to induced, but rather seeks to better understand reproductive health as a whole among refugees. We made this decision because all reproductive health services are linked, and levels of gender-based violence and contraceptive use, for example, affect abortion rates, as does access to clinical management of rape services and family planning services.

##### Search criteria

On 22 September 2023, a scoping review was conducted for the last five years ( starting 23 September 2018 and ending on the search date) of English language peer-reviewed journal articles using PubMed and Google Scholar. We selected PubMed due to its vast collection of life sciences and biomedical research. We used Google Scholar as a secondary database in our search due to its expansive collection of research, including items published outside of typical health science journals. We tested several combinations of search terms and synonyms, and ultimately decided to search using the following terms after determining that they produced the most relevant results: “(refugee OR displaced OR displacement) AND (“reproductive health” OR “sexual health” OR fertility OR contraception OR abortion)”. We reviewed all PubMed results in the last five years and the first 300 Google Scholar results sorted by relevance. We decided to review only the first 300 Google Scholar results due to their accessibility and relevance, and to increase the efficiency of our search. We chose these criteria to best identify emerging or evolving topics and gaps related to refugee fertility intentions and access to reproductive health services.

##### Analysis

We downloaded search results into Zotero citation management software^34^ and exported the records into Microsoft Excel for screening. Once we arrived at the final sample based on items’ inclusion of results related to refugee fertility intentions and access to reproductive health, we moved the data into Microsoft Word. We identified themes in the data through discussion and paying particular intention to fertility intentions and how displacement affected actual fertility and intended fertility. We then developed summaries based on themes we identified in the data. We did not exclude individual research articles that were included in review articles that we included in our final sample. We did not conduct any critical appraisal of the items included in the scoping review. We report all literature review results according to PRISMA for Scoping Reviews 22-item checklist.^35^ The scoping review protocol is not published but is available from the authors upon request.

## Results

### Quantitative results

We downloaded the records of 29,413,033 individuals from the UNHCR Refugee Data Finder for 2022. We eliminated individuals of unknown sex (n=3,745,164), male sex (12,501,718), female age unknown (2,025,378), and female age <12 years and 60+ years (n=4,184,188), leaving a population of 6,956,585 individuals. We then removed individuals with origin countries of China, Hong Kong SAR (n=41), Western Sahara (n=83), unknown (n=7,839), and stateless (n=2,918). We also removed individuals with host country China, Hong Kong SAR (n=83). Finally, after merging these data with information from the Center for Reproductive Rights and the Council on Foreign Relations, we eliminated individuals with origin country or host country Colombia (n=30,196 and n=626), Thailand (n=46 and n=34,201), and the United States (n=96 and n=20,270) due to law changes in 2022. Our final sample includes data from 6,860,398 female refugees aged 12-59 years who sought asylum in 2022. A flow diagram in *Figure 1* shows how we narrowed down the population included in our analysis.

**Figure 1.**
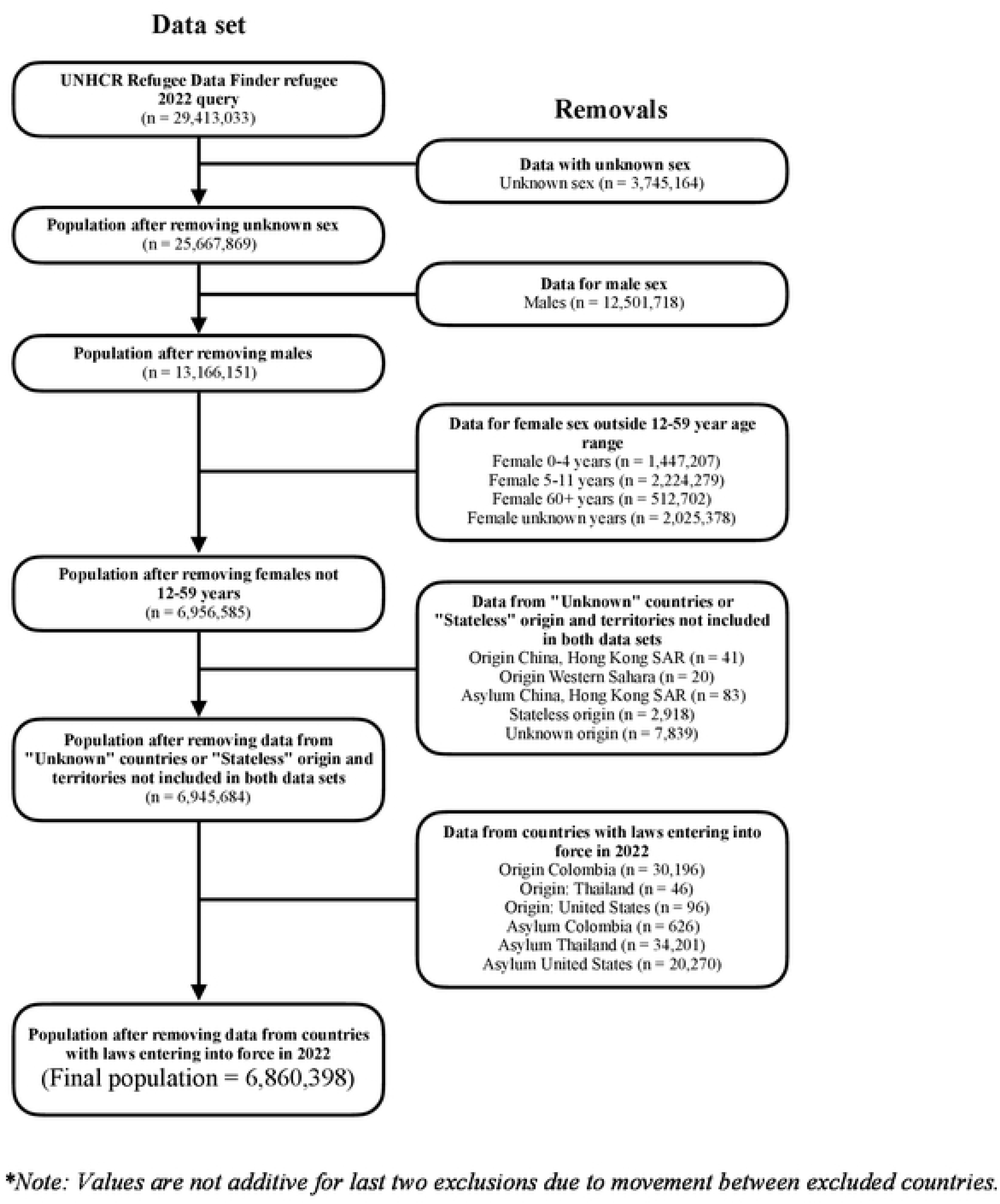
Flow diagram of study population inclusion and exclusion*

The final data set included 159 origin countries and 111 host countries, though it should be noted that the UNHCR Refugee Data Finder combines Serbia and Kosovo into one entity. Refugees originated from 53 countries that were not destinations for refugees; and five host countries did not generate refugees. *S1 Appendix* is a list of the origin and host countries included in the analysis and the total number of refugees originating in and seeking asylum in each. Heat maps in *Figure* 2 show the geospatial distribution of individuals originating and seeking asylum in each country. While 26% of refugees went to countries with abortion on request, 5% went to countries with abortion for socioeconomic reasons, 28% went to countries with abortion to preserve health, 38% went to countries with abortion to save the pregnant person’s life, and 3% went to countries where abortion is prohibited altogether. The Syrian Arab Rep. generated the most female refugees aged 12-59 years old in 2022 (26% of total sample) who sought asylum in 64 countries. Iran (Islamic Rep. of) accepted the most female refugees aged 12-59 years old in 2022 (15% of total sample), who originated from Afghanistan, Iraq, Kuwait, and Pakistan. The largest flows of female refugees aged 12-59 years old were from Afghanistan to Iran (Islamic Rep. of) (15% of total sample), Syrian Arab Rep. to Turkïye (14% of total sample), Ukraine to Poland (6% of total sample), Afghanistan to Pakistan (6% of total sample), and Ukraine to Germany (5% of total sample).

**Figure 2.**
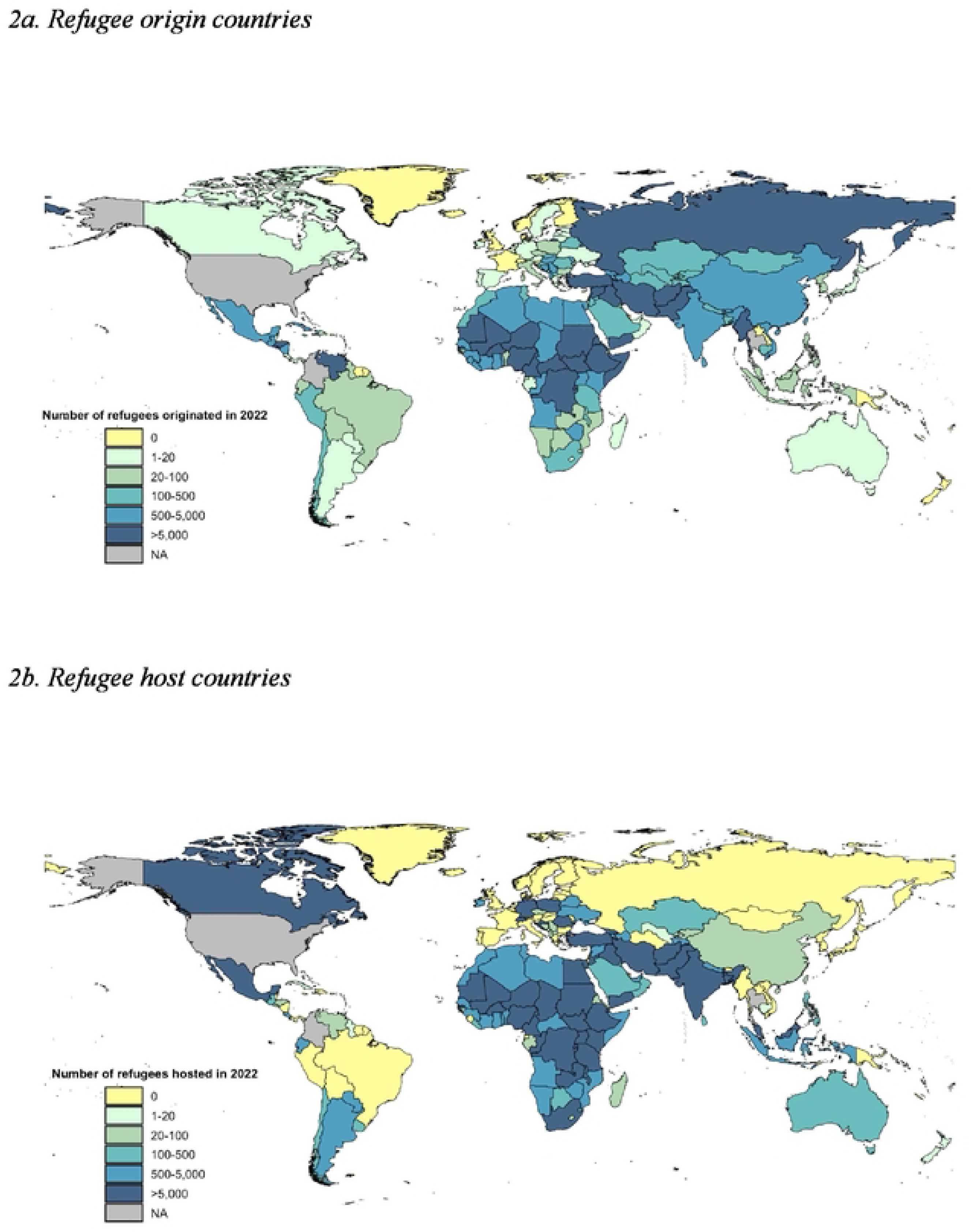
Global heat map showing refugee origin and host countries in 2022, for female refugees aged 12-59 years

#### Travel from countries with less restrictive laws to countries with more restrictive laws

1,083,677 female refugees aged 12-59 years old (16%) sought asylum in countries with more restrictive laws than their origin country in 2022. These refugees originated in 61 counties and sought asylum in 60 countries. The most commonly represented abortion law category for refugee origin was 3, countries with abortion to preserve health, which were involved in 121 or 44% of exchanges in 2022; 61 (22%) of exchanges originated in countries with abortion on request, category 1, 46 (17%) in countries with abortion on socioeconomic grounds, category 2, and 49 (18%) in countries with abortion to save the pregnant person’s life, category 4.

Conversely, the most commonly represented abortion law category for refugee asylum was 4, countries with abortion to save the pregnant person’s life, which were involved in 117 or 42% of exchanges in 2022; 6 (2%) of exchanges originated in countries with abortion on socioeconomic grounds, category 2, 50 (18%) in countries with abortion on to preserve health, category 3, and 106 (38%) in countries with abortion prohibited altogether, category 5. The largest contributors to this group were refugees from Ukraine fleeing to Poland (n=413,166), from the Dem. Rep. of the Congo fleeing to Uganda (n=136,342), and from the Syrian Arab Rep. fleeing to Iraq (n=73,583). *Figure 3* shows the exchange of refugees between countries with less restrictive to more restrictive abortion laws, with refugees moving from the lefthand column to the righthand column.

**Figure 3.**
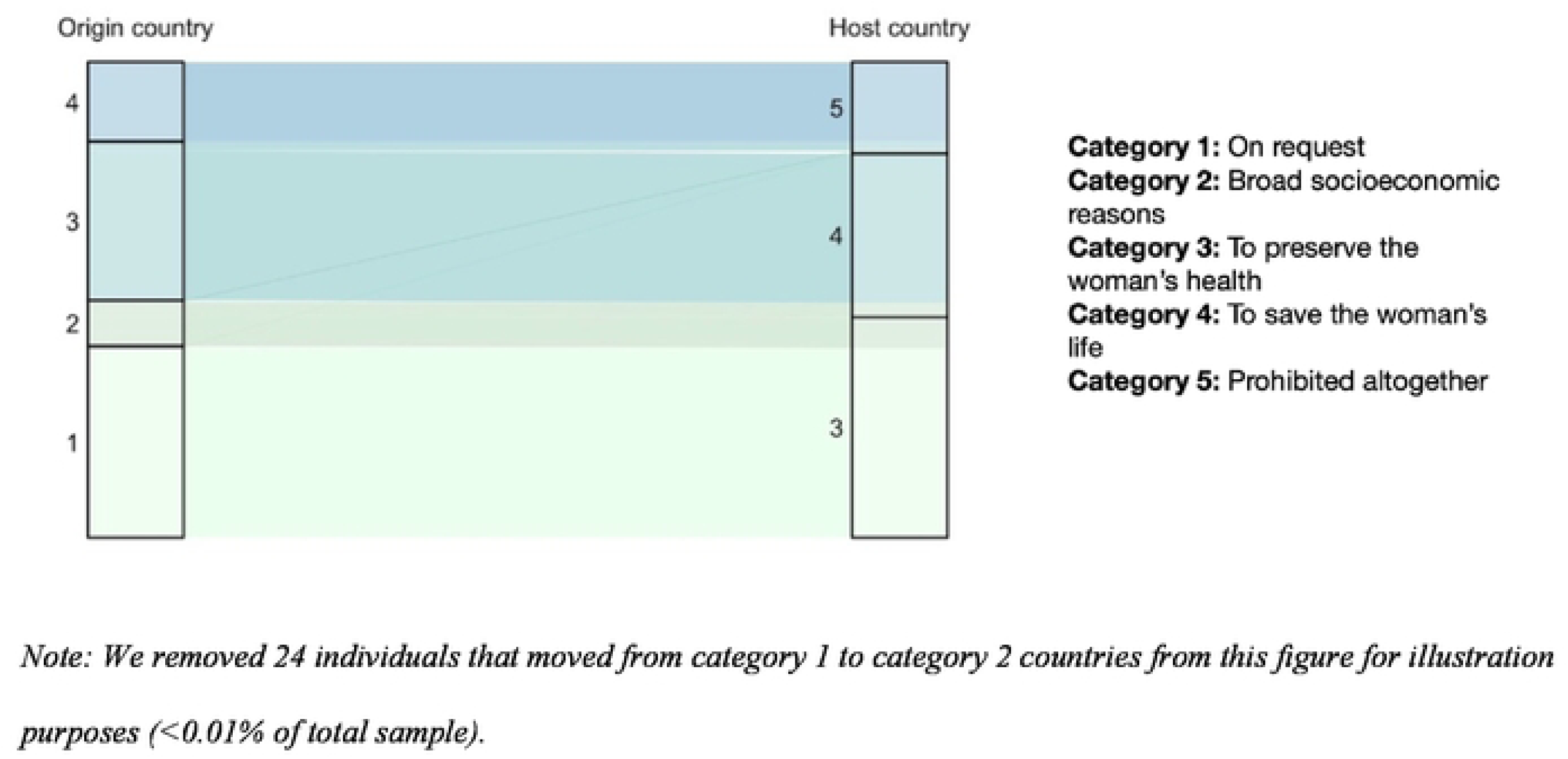
Segment of refugee population moving between countries with less restrictive to more restrictive abortion laws based on abortion law category, 2022*

#### Travel between countries with the same assigned abortion law category

3,003,005 female refugees aged 12-59 years old (44%) sought asylum in countries with abortion laws in the same abortion law category as their origin country in 2022. These refugees originated in 109 counties and sought asylum in 90 countries. By far, the most commonly represented abortion law category for each country pair was 1, countries with abortion on request, which were involved in 205 or 48% of exchanges in 2022; only 5 (1%) of exchanges originated in countries with abortion on socioeconomic grounds, category 2, 106 (25%) in countries with abortion to preserve health, category 3, and 92 (22%) in countries with abortion to save the pregnant person’s life, category 4, and 17 (4%) in countries with abortion prohibited altogether. The largest contributors to this group were refugees from Afghanistan fleeing to Iran (Islamic Rep. of) (n=1,013,085), from Ukraine fleeing to Germany (n=317,636), and from Myanmar fleeing to Bangladesh (n=291,481).

#### Travel from countries with more restrictive laws to countries with less restrictive laws

In 2022, 2,774,716 female refugees aged 12-59 years old (40%) sought asylum in countries with more restrictive laws than their origin country. These refugees originated in 98 counties and sought asylum in 87 countries. Interesting, although this category accounted for the second most individuals who fled from one country to another in 2022, it included the most exchanges between countries: 702. The most commonly represented abortion law category for refugee origin was 4, countries with abortion to save the pregnant person’s life, which were involved in half, or 352 (50%) of exchanges in 2022; 28 (4%) in countries with abortion on socioeconomic grounds, category 2, 178 (25%) in countries with abortion to preserve health, category 3, and 144 (21%) in countries with abortion prohibited altogether, category 5. Conversely, the most commonly represented abortion law category for refugee asylum was 4, countries with abortion to save the pregnant person’s life, which were involved in 117 or 42% of exchanges in 2022; 1 (2%) of exchanges originated in countries with on socioeconomic grounds, category 2, 50 (18%) in countries with abortion on to preserve health, category 3, and 106 (38%) in countries with abortion prohibited altogether, category 5. The largest contributors to this group were refugees from the Syrian Arab Rep. fleeing to Turkïye (n=986,895), from Afghanistan fleeing to Pakistan (n=413,174), and from the Syrian Arab Rep. fleeing to Jordan (n=206,072). *Figure 4* shows the exchange of refugees between countries with more restrictive to less restrictive abortion laws, with refugees moving from the lefthand column to the righthand column.

**Figure 4.**
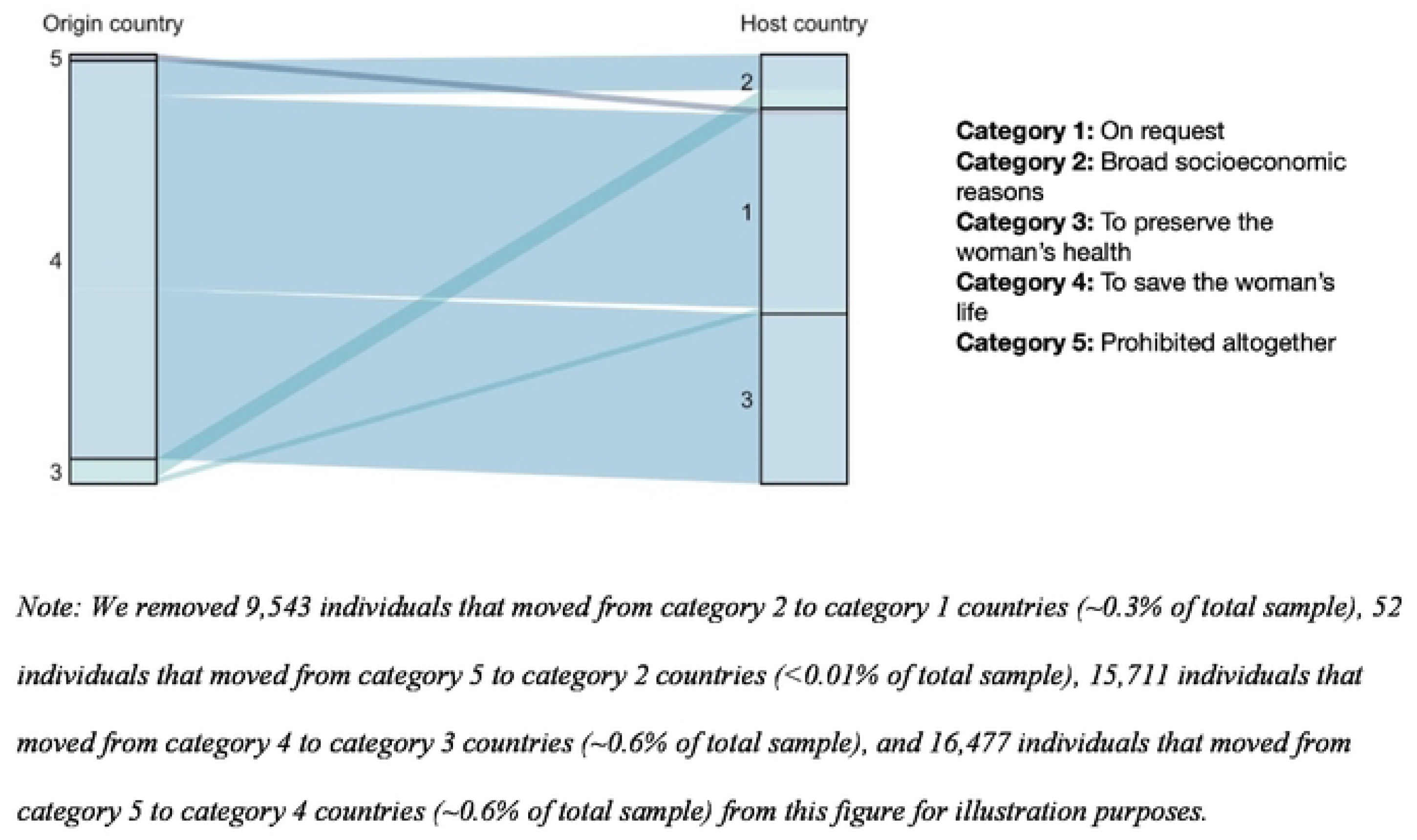
Segment of refugee population moving between countries with more restrictive to less restrictive abortion law categories, 2022*

### Scoping review

The results of our scoping review contextualize the findings from our quantitative analysis. Our search resulted in 566 results on PubMed and 26,100 on Google Scholar, of which we extracted 300, leading to a total of 866 items. We removed 73 duplicates, resulting in 793 unique items. We next removed items published in a language other than English (n=3). After removing books and book sections (n=15), dissertations and theses (n=9), commentaries (n=7), abstracts (n=6), and other non-peer reviewed journal articles (n=11), we were left with 742 items to screen. We then removed 361 items that were not relevant to sexual and reproductive health, 43 items that did not focus on refugees, and 286 items that did not provide data to answer our research questions. Fifty-two (52) items are included in the synthesis below. Our screening process is charted in a flow diagram in *Figure 5.* The majority of articles did not address our research questions directly, but rather explored refugee use of various reproductive health services. The most common countries of origin for items were “Any” (n=11), Myanmar (n=10), and Syria (n=10), though fifteen countries were named. The most common host countries were the United States (n=8), Bangladesh (n=7), Uganda (n=7), though fifteen countries were named. Thirty-two (32) items explored fertility intentions and 29 reported on access to sexual and reproductive health services. More characteristics of each item included in the synthesis are available in *S2 Appendix*.

**Figure 5.**
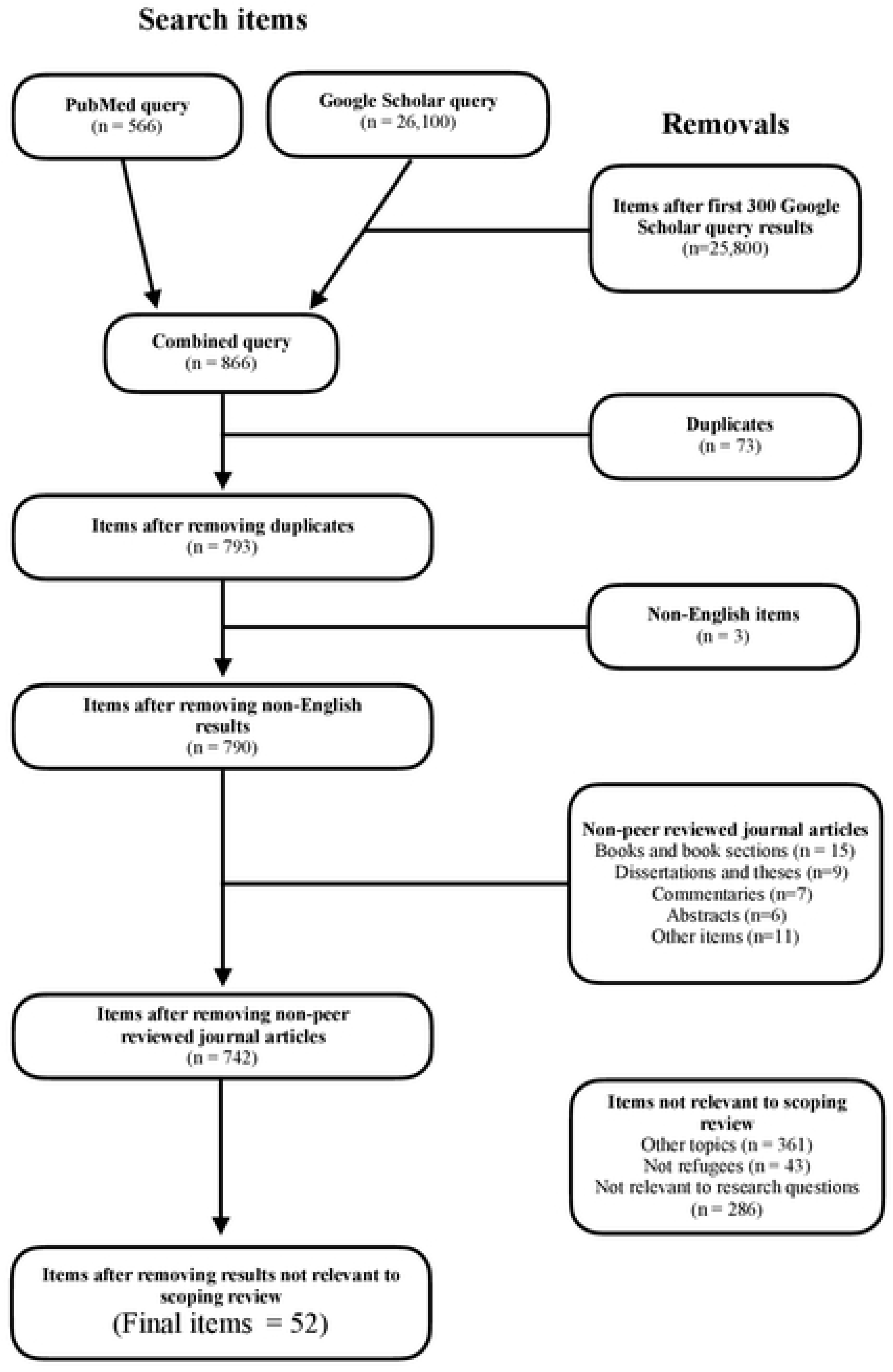
Flow diagram of scoping review inclusion and exclusion

While there is overlap in the themes presented in each article, we determined that there were distinct themes related to refugee health and present our results in the following sections: (1) unintended fertility; intended fertility; need to space, postpone, and/or limit births; changing fertility intentions; and (2) access to reproductive health services. In intended and unintended fertility sections, we discuss how refugee status affects individuals’ abilities to regulate their fertility; in the need to space, postpone, and/or limit births section, we summarize how forced displacement affects fertility desires; and in the changing fertility intentions, we analyze the factors that cause the changes in desires around spacing, postponing, and limiting births. In the access to reproductive health services section, we describe how refugee status affects service provision and receipt.

Most research published in the past five years on fertility intentions and pregnancy among refugees focused on adolescents and youth. These studies found that in most cultures, sex and pregnancy before marriage including unintended pregnancies due to rape were stigmatized and had severe consequences (unintended fertility), but younger age at marriage and subsequent childrearing were encouraged (intended fertility).

#### Unintended fertility

Several studies explored the specific vulnerabilities facing adolescent refugees that exposure them to unintended pregnancies. A qualitative study among pregnant teenagers and teenage moms in Mahama refugee camp in Rwanda found that premarital sex was seen as proof of “bad behavior” due to its association with pregnancy and HIV/AIDS, and that pregnant teenagers are seen as “bad influences” who made deliberate decisions to become pregnant, even when their pregnancies are the result of sexual assault.^36^ Focus group discussions with 12-17-year old Congolese refugees in Rwanda revealed that unintended pregnancies were often the result of financial insecurity, which drove girls to exchange money for sex or goods with individuals outside of the refugee camps where they lived.^37^ A cross sectional study among various adolescent girl refugees in Uganda found that they were scared to become pregnant before marriage, which could lead to social exclusion and expulsion from school, but that coerced, forced, and transactional sex were common in the camps, as were coerced and forced births by partners.^38^ A study examining maternal health service seeking behavior similarly reported that many pregnancies among refugee youth in Uganda were due to rape and not desired fertility.^39^ Coercive and transactional sex among adolescent and youth refugee and host community members in Rwanda made them vulnerable to unintended pregnancies.^40^ Adolescent Rohingya migrant and refugee boys and girls in Thailand believed they were too young for children and worried about educational, social, and financial consequences in addition to adverse health impacts.^41^ More than half of study participants reported that their current pregnancies were unintended. Two studies among youth migrants and refugees in Australia reported that unintended pregnancies were stigmatized, and could lead to forced marriage and mistreatment, among other social consequences.^42,43^ While not limited to adolescents, a third Australian study found that reproductive coercion led to violence during pregnancy, including forced miscarriage, abortion, pregnancy, and contraceptive sabotage among refugees and migrants.^44^ This was due to extra-marital sex being taboo despite cultural pressure to have large families.

#### Intended fertility

Child marriage in refugee settings is related to community desires for large families. Key informant interviews with Syrian refugees in Egypt explored child marriage and emphasized the importance of childbearing from a young age, despite health repercussions for young mothers.^45^ Similarly, a study on child marriage among Rohingya refugees in Bangladesh found that cultural preferences for adolescent brides, prevailing social norms, and insecurity led to these marriages and subsequent childbearing at a young age.^46^ A mixed methods study in Bidi Bidi refugee settlement also found that individuals experiencing forced child marriage were significantly more likely to report forced pregnancy, abortion, and missed educational opportunities due to sexual violence.^47^

Studies among married adults found that pregnancy and childbirth were often seen as positives. A qualitative study among Rohingya refugees in Bangladesh reported that having a more children was viewed positively in the community because those children would continue the couple’s lineage and contribute to the Islamic population in Bangladesh.^48^ While Rohingya women had access to contraceptives, they did not want their marriages to end because they were unwilling to have (more) children with their spouses. Rohingya women refugees in another study in the same context reported the same, that large families were preferred in their culture and that contraceptive use was stigmatized in the culture and religion.^49^ In this study, however, participants did share that this view was actively shifting. A third study among Rohingya refugees found that 48% of female respondents did not participate in fertility decision-making and that this non-participation and low participation were significantly associated with shorter birth intervals.^50^ A qualitative study in Canada and Australia among refugees from countries in South America, Asia, and Africa had similar findings: participants discussed children as a central part of a woman’s identity and imperatives for their culture and religion.^51^ A systematic review of Syrian refugee fertility in Turkïye revealed that high fertility continued after resettlement, but was driven by husbands and not mothers,^52^ and a study among refugee women in the country confirmed the desire for higher fertility: participants reported an average of 2.37 living children, with a desire for an average of 3.79 children.^53^ One study in Bangladesh revealed that a majority of Rohingya refugee women (58%) thought childbearing should continue until a son is born.^54^ Another study in the same population found that religion, son preference, and community pressure were common drivers of high fertility.^55^

#### Need to space, postpone and/or limit fertility

Some studies reported that refugee status made women want fewer children due to decreased social supports and environmental constraints. In a qualitative study among Somalian and Congolese refugees in the United States, participants discussed the cultural and religious pressure to have large families, but also the increased difficulty of raising children after displacement due to social isolation, and the loss of multigenerational families and traditions that come with a community.^56^ Congolese refugees stated that these were significant factors in their fertility desires. A series of focus groups conducted among Somali refugees in Kenyan refugee camps found similarly contradictory views -- participants discussed the benefits of large families in their culture, such as additional income, caregiving, and esteem in the community, and specific to their current situation, reporting that more children meant extra ration cards and food.^57^ At the same time, participants spoke about the adverse health risks associated with young and frequent childbearing, in addition to added responsibilities and fewer economic and educational opportunities. Another paper exploring Somali refugee family planning use in the United States reported similar tensions between fertility desires and constraints.^58^ Focus group participants felt pressure from their community to conceive after marriage, and emphasized the role of God in planning for family size and timing. However, at the same time, participants shared that they wanted to space births, but did not feel that they had control to do so, and that the lack of family support in the United States was an incentive to use contraception following childbirth. A similar study using focus groups in the same population found that couples often differed on family size, with wives pushing for smaller families and husbands requesting more children.^59^

Other studies found mixed sentiments among adult refugee women regarding their fertility. A mixed methods study examining family planning use among South Sudanese refugees and host communities in Uganda reported that participants were caught in tension between their desire to have large(r) families and their ability to provide for that family.^60^ Participants described in detail their struggles to farm on land granted to them by the Ugandan government and issues with finances that impacted their fertility desires. A study among Syrian refugees in Lebanon found that political, economic, and health crises in Lebanon actively influenced fertility desires of refugees: decreased social support, deteriorated living conditions, and lack of jobs led to Syrian refugee women wanting to limit their births.^61^ Among Bhutanese, Rohingya, and Iraqi refugees in the United States, participants reported that the stability associated with moving from a fraught environment empowered women, which included increasing their desire for family planning.^62^ A survey of 300 Syrian refugees in Turkïye found that the average family size in the study group was less than the ideal family size; 37% of respondents reported having less than two-year pregnancy intervals and 43.6% reported having had their last child against their will.^63^ Only one study among adults explored forced sex: a study among asylum-seekers and refugees in South Africa reported that many individuals were victims of forced sex and rape during their migration journey or felt that they needed to engage in transactional sex for survival.^64^

#### Changing fertility intentions

Two review articles and a study in Lebanon revealed the factors affecting fertility decision-making before displacement, during displacement, and after arrival. In a scoping review and thematic analysis of influences on fertility among refugees in high-income countries, these factors included things like religious beliefs, active conflict, and coercion.^65^ Another systematic review similarly explored the effect of refugee status on contraceptive care in high-income countries.^66^ This study reported that many refugees originated in countries with cultural expectations of large families, and some individuals felt pressured to continue this tradition to recreate the security and feeling of former communities, while for some, resettlement led to changes in fertility and contraceptive desires to align with a desire for smaller families. A comparison of parity among Syrian refugees in Lebanon who became pregnant before, during, and after displacement found that for individuals who got pregnant in Syria only, there was median gravidity of five to six, but among refugees only pregnant in Lebanon, after displacement, median gravidity was two.^67^ Interestingly, individuals getting pregnant in Lebanon exclusively were younger and significantly more likely to access abortion services.

#### Access to reproductive health services

Access to reproductive health services for refugees is constrained across the socioecological model, as demonstrated by research published in the past five years. A summary of the different factors mentioned in publications at their respective locations in the socioecological model is available below in *Figure 6*, which was developed by the authors as a way to make the interpretation of these findings clearer, so that they are actionable to stakeholders who may be considering interventions at each level.

**Figure 6.**
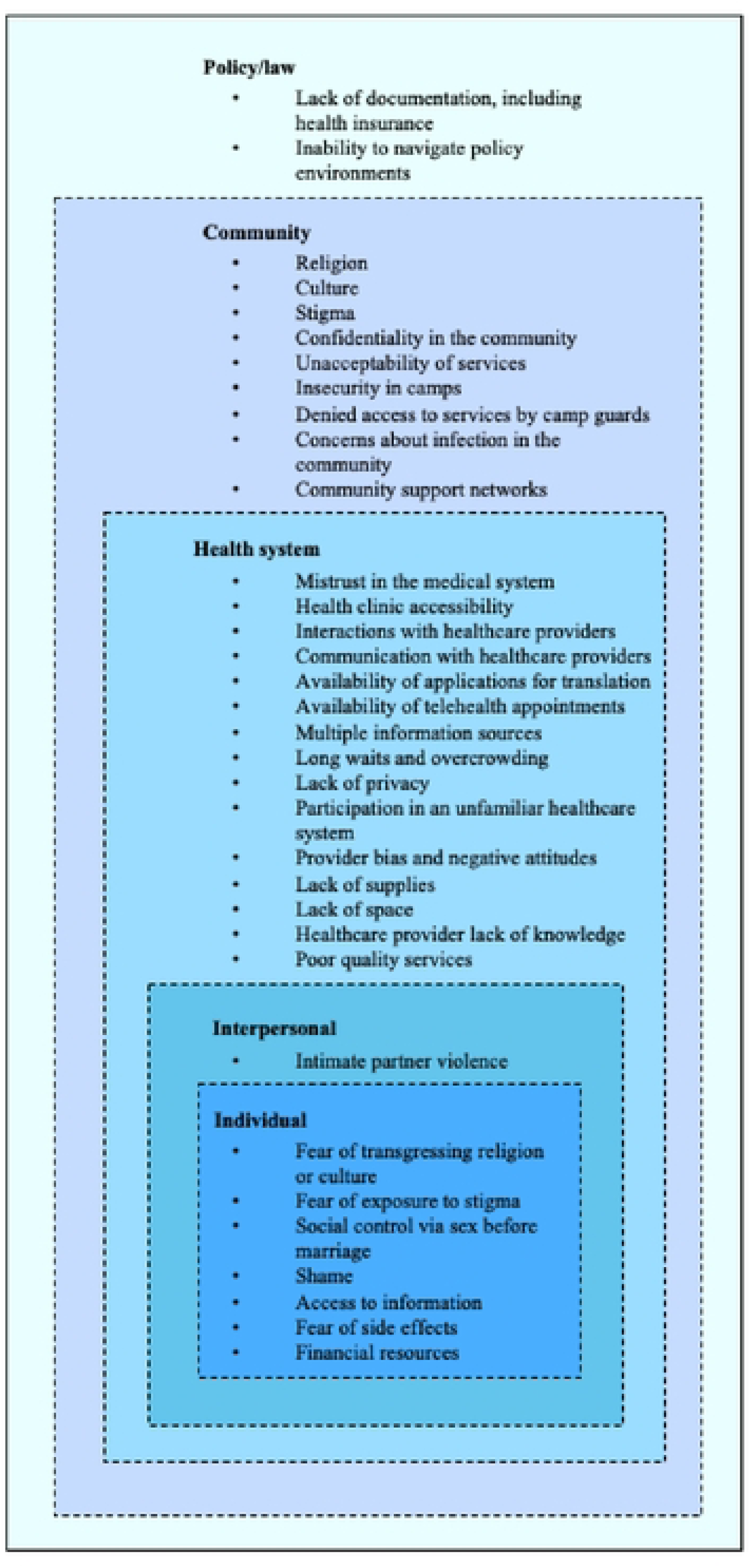
Factors affecting refugee access to reproductive health services at different levels of the socioecological model

At the individual level, studies show that access to reproductive health knowledge is made hard to access by fears of transgressing religion or culture, and exposure to stigma, especially for adolescents.^68^ Among adolescents in Bangladesh, fear of stigma and social control via sex before marriage prevented them from accessing reproductive health services.^69^ Shame among individuals who had been sexual assaulted prevented them from accessing health services, and the same study reported that adolescents faced even more burdens, especially when unmarried.^70^ Though sometimes displacement and resettlement increases access to information,^71^ a systematic review of refugee research globally and two studies including interviews with adolescent refugees in Uganda and Bangladesh revealed that many participants did not know where to seek reproductive health services.^46,72,73^ Fears of side effects from contraceptives may also impede access to services at the individual level.^48^ A lack of financial resources was the most commonly discussed barrier to services for refugees.^39,64,66,72–80^

Very few factors at the interpersonal level of the socioecological model were discussed in items included in our scoping review. Husbands, and especially the threat of intimate partner violence, were noted as significant barriers to reproductive health access, including and especially, contraceptive access.^48^ Internally displaced persons and refugees in Kenya reported that they did not seek out sexual- and gender-based violence (SGBV) treatment due to fear of future abuse.^81^ Research items did not discuss other individual relationships and the role they played in access to services.

At the health system level, fears of contraception may be exacerbated by mistrust in the medical system.^48^ Qualitative studies in the United States and other countries found that facilitators to services included accessible clinics and positive interactions with healthcare providers,^74^ while an inability to communicate with healthcare providers created a barrier to services.^39,52,66,73,74,78,82^ Mobile applications for translation, telehealth appointments, and multiple information sources helped to facilitate access to services where individuals faced communication barriers.^73^ South Sudanese refugees in Uganda and cited distance, long waits, privacy, and overcrowding as the main barriers to accessing reproductive health services, though 63.3% of surveyed individuals felt that health facilities in refugee settlements were satisfactory overall.^60^ A study among adult refugees in high-income countries found similar accessibility issues, like difficulty using public transportation and participating in an unfamiliar healthcare system.^66^ Provider bias and negative attitudes affected refugees in Rwanda,^40^ Uganda,^39,83^ Israel,^79^ and South Africa^64,78^ in their plight to access services. A systematic review pointed to healthcare provider training on cultural sensitivity and communication as a facilitator of access.^73^ Lack of supplies also acted as a barrier in some places, like for Syrian refugees in Lebanon,^77^ Eritreans in Israel,^79^ and Congolese individuals in Uganda.^80^ However, even where supplies were available, like in Bangladesh for Rohingya refugees, a lack of space prevented service provision.^69^ And, even in locations with less restrictive abortion laws, healthcare providers’ lack of knowledge presented additional barriers to services for refugees.^69^ Even when healthcare providers were able to give reproductive health services, refugees in Uganda and Israel thought they were unacceptable and/or poor quality.^79,83^

Adolescent refugees cited these same issues at the health systems level and more. Long waits at health facilities were also by adolescent Rohingya refugees in Bangladesh,^46^ Syrian refugees in Lebanon,^84^ and Eritrean refugees in Israel,^79^ as were overcrowded facilities for refugees in Uganda,^39^ and both were reported as issues in South Africa.^78^ Adolescent refugees surveyed in Uganda and Syria did not return to health facilities for reproductive health services after an initial visit due to lack of privacy, resources, distance to facilities, and mistreatment by healthcare providers.^38,84^ A study among Rohingya refugees in Bangladesh, confirmed that discrimination, including healthcare providers refusing to give contraceptives to adolescents, prevented them from using family planning.^41^ Refugees in Uganda likewise complained about lack of adolescent access to health services and information.^83^

At the community level, religion, culture, and stigma prevent access to reproductive health services.^68,78,85^ A systematic review reported that adolescents were concerned about confidentiality due to stigma associated with sexually transmitted infections and premarital sex.^72^ A systematic review of barriers and facilitators of access for migrant, internally displaced, refugee, and asylum-seeking women cited culturally unacceptability of services as a major deterrent.^73^ Among refugees and internally displaced persons in the Dadaab refugee complex in Kenya, there were several community-level barriers to care following SGBV, including stigma in the community, insecurity in the camps, and being denied access by guards.^81^ During the COVID-19 pandemic, Syrian refugees in Lebanon had trouble accessing services due to concerns about infection and increased insecurity.^75^ Community support networks helped to mitigate these barriers in Kenya^81^ and South Africa.^78^

At the policy/law level, lack of documentation,^52,64^ including health insurance,^76,82^ may act as an impediment to health services. In only one study, among Congolese refugees in Uganda, participants reported being unable to navigate the abortion policy environment,^86^ where abortion is illegal except to save the pregnant individual’s life.^87^ This lack of navigability led refugees to access less safe abortion procedures and left them unable to access post-abortion care for related complications.

## Discussion

The results of this analysis begin to fill an important gap in understanding how forcible displacement may affect refugees’ fertility decision-making, their ability to control their fertility and access reproductive health services, and how legal landscapes may affect their choices and access. Our quantitative analysis of UNHCR and CRR data revealed that nearly one in five female refugees aged 12-59 years old moved from countries with less restrictive to more restrictive abortion laws and that only 24% of these refugees were hosted in countries with abortion on request.

Though key humanitarian organizations, including Doctors without Borders, have implemented safe abortion care in locations that they work, including refugee camps, many humanitarian organizations fear doing so due to legal repurcussions.^88–90^ The organizations that do provide these services interpret abortion rights for refugees to be protected by international humanitarian law that considers pregnant individuals to be “wounded and sick”, entitled to medical care “to the fullest extent practical”, with “least possible delay”, and “no adverse distinction”, i.e., discrimination due to female sex.^90^ In line with this interpretation, care should be provided whether or not the need for care, i.e., pregnancy, arises from something related to the conflict (or displacement) or not. Various United Nations treaties also protect the right of refugees to safe abortion and post-abortion care.^15^

An additional barrier to health services not addressed in our quantitative analysis or scoping review is that of status designation in host countries. For example, while the UNHCR designates these individuals as refugees, the Turkïye government gives Syrians “temporary protection status,” and in Bangladesh, the government calls Myanmar refugees “Forcibly Displaced Myanmar Nationals”.^91^ These and other host nations’ practices around designating individuals as “refugees” versus other entities, regardless of UNHCR designation, negatively affects their ability to access health services.^92^ This analysis draws attention to the need for more funding, guidance, and support for entities to provide access to abortion services to refugees, as defined by the UNHCR, wherever they implement other reproductive health services.

The results of the scoping review complement and contextualize the findings of our quantitative analysis. They point to the effect that refugee status has on fertility intentions: adult female refugees expressed mixed fertility intentions, but adolescents, especially unmarried adolescents, wanted to control their reproduction, but were left unable to in most cases due to community stigma, restricted access to family planning services, and high levels of sexual- and gender-based violence in refugee settings. Studies included in our synthesis demonstrated the commonality of sexual coercion, transactional, and forced sex within refugee settings. A meta-analysis conducted among female refugees in complex humanitarian emergencies confirms this result, finding that the estimated prevalence of sexual violence in refugee populations is 21.4%.^8^ Other studies in the scoping review among adolescents found that forced child marriage was common due to the cultural desire for larger families. A study in 34 African countries reported that children married before the age of eighteen years were more likely to experience rapid childbirth and larger family sizes than individuals married at the age of majority or older.^93^ The story was less clear among adults — some studies found that refugees wanted large families regardless of refugee studies, others reported that refugee status made individuals want fewer children due to lack of social support networks, and a third group of studies reported that individuals experienced tensions between the two sentiments. While few studies exist on the topic, research in Zimbabwe found that individuals believed that community support was necessary for healthy pregnancies and child-rearing.^94^ Individuals everywhere should be supported in their decision-making around their own fertility.

The scoping review demonstrated the multifaceted constraints that refugees, and especially adolescent refugees, face in accessing high-quality reproductive health services. Most studies documented these barriers at the community, health system, and individual level, while a few studies reported additional barriers at the interpersonal and policy/law level of the socioecological model. Very few items included in our scoping review found facilitators for reproductive health services among refugees. This is not surprising: quantitative analyses have found that up to 19.9% of hospital admissions in fragile settings^95^ and 25-50% of maternal deaths in refugee settings are due to abortion complications.^96^ These studies’ quantitative findings, coupled with the results in our scoping review, and our own quantitative analysis point to the imperative that refugees have access to high-quality health services across their reproductive life course.

### Limitations

There are several limitations of this analysis. We limited our quantitative analysis, and our qualitative synthesis, where possible, to only include refugees and not other forcibly displaced populations. We did this because fertility intentions and access to reproductive healthcare, including abortion, may differ between different groups of forcibly displaced populations. However, we acknowledge that there is heterogeneity even within refugee populations which we do not explore, given our goal to characterize the topic globally. In our quantitative analysis, we were constrained by the limits of our data sets: while “women of reproductive age” are considered to be 15-49-year-olds, we were only able to subset the entire population of refugees to females aged 12-59 years. In addition, we removed countries whose abortion laws changes during 2022 because we only had data aggregated to an annual time period. Our scoping review is limited by our decision to include only publications in the English language, that are indexed to PubMed and Google Scholar, published in the last five years, and related to our chosen search terms. We made these decisions to include the most relevant research to characterize the field at this moment in time. Despite these limitations, we believe that this is an important and novel analysis that contributes to the field’s better understanding of how individual’s abortion rights are affected by forced migration, and what research in the last five years has revealed about refugee fertility intentions and access to reproductive health services.

## Conclusions

All individuals capable of pregnancy, whether forcibly displaced or part of another population, deserve the ability to control their reproduction. This study reveals the various constraints that refugees may face in their attempts to make decisions about and control their fertility, and to access reproductive health services, including safe abortion and post-abortion care. The data also point specifically to the need to continue to protect refugee girls from early marriage, early childhood, and unintended pregnancies. The results of the quantitative analysis and scoping review demonstrate the need for a renewed international commitment to ensuring that a key vulnerable population, refugees, can access the full range of high-quality reproductive health services to so they’re able to decide if, when, and how many children to have.

*S1 Appendix. Countries included in the analysis, their law category, and number of refugees originated and seeking asylum within their borders*

*S2 Appendix. Items included in the scoping review*

## Data Availability

All data generated or analyzed during this study are included in this published article or are publicly available online.

## List of abbreviations

CAT: Convention against Torture and Other Cruel, Inhuman or Degrading Treatment or Punishment
CEDAW: Convention on the Elimination of All Forms of Discrimination against Women
CRR: Center for Reproductive Rights
SGBV: Sexual- and gender-based violence
UDHR: Universal Declaration of Human Rights
UNHCR: United Nations High Commissioner for Refugees
WRA: Women of reproductive age

## Acknowledgements

Not applicable

## Declarations

### Ethics approval and consent to participate

Not applicable.

### Consent for publication

Not applicable.

### Authors’ contributions

- Blake Erhardt-Ohren: conceptualization, data curation, formal analysis, investigation, methodology, project administration, software, visualization, writing - original draft
- Ndola Prata: conceptualization, investigation, methodology, project administration, supervision, validation, writing - review & editing

## References

1. UNHCR Global Trends - Forced displacement in 2020. UNHCR Flagship Reports. Accessed May 10, 2022. https://www.unhcr.org/flagship-reports/globaltrends/

2. Refugees UNHC for. UNHCR - Refugee Statistics. UNHCR. Accessed April 19, 2023. https://www.unhcr.org/refugee-statistics/

3. Refugees UNHC for. Climate change and disaster displacement. UNHCR. Accessed May 11, 2022. https://www.unhcr.org/climate-change-and-disasters.html

4. Briefing paper: Understanding the root causes of displacement, IDMC 2015. Accessed May 10, 2022. https://www.unhcr.org/protection/operations/56684ce89/briefing-paper-understanding-root-causes-displacement-idmc-2015.html

5. Tanabe M, Myers A, Bhandari P, Cornier N, Doraiswamy S, Krause S. Family planning in refugee settings: findings and actions from a multi-country study. Confl Health. 2017;11(1):9. doi:10.1186/s13031-017-0112-2

6. Volkov VG, Granatovich NN, Survillo EV, Pichugina LV, Achilgova ZS. Abortion in the Structure of Causes of Maternal Mortality. Rev Bras Ginecol Obstet. 2018;40(6):309–312. doi:10.1055/s-0038-1657765

7. Ward J, Vann B. Gender-based violence in refugee settings. The Lancet. 2002;360:s13–s14. doi:10.1016/S0140-6736(02)11802-2

8. Vu A, Adam A, Wirtz A, et al. The Prevalence of Sexual Violence among Female Refugees in Complex Humanitarian Emergencies: a Systematic Review and Meta-analysis. PLoS Curr. 2014;6:ecurrents.dis.835f10778fd80ae031aac12d3b533ca7. doi:10.1371/currents.dis.835f10778fd80ae031aac12d3b533ca7

9. Sardinha L, Maheu-Giroux M, Stöckl H, Meyer SR, García-Moreno C. Global, regional, and national prevalence estimates of physical or sexual, or both, intimate partner violence against women in 2018. The Lancet. 2022;399(10327):803–813. doi:10.1016/S0140-6736(21)02664-7

10. Dias Amaral B, Sakellariou D. Maternal Health in Crisis: A Scoping Review of Barriers and Facilitators to Safe Abortion Care in Humanitarian Crises. Frontiers in Global Women’s Health. 2021;2. Accessed May 10, 2022. https://www.frontiersin.org/article/10.3389/fgwh.2021.699121

11. Abiola AH, Oke O, Balogun M, Olatona F, Adegbesan-Omilabu M. Knowledge, attitude, and practice of abortion among female students of two public senior secondary schools in Lagos Mainland Local Government Area, Lagos State. Journal of Clinical Sciences. 2016;13(2):82–87. doi:10.4103/2408-7408.179682

12. Klc TR, Ames S, Zollinger B, et al. Abortion in rural Ghana: Cultural norms, knowledge and attitudes. African journal of reproductive health. 2020;24(3):51–58. doi:10.29063/ajrh2020/v24i3.6

13. Makleff S, Labandera A, Chiribao F, et al. Experience obtaining legal abortion in Uruguay: knowledge, attitudes, and stigma among abortion clients. BMC Women’s Health. 2019;19(1):155–155. doi:10.1186/s12905-019-0855-6

14. Munakampe MN, Zulu JM, Michelo C. Contraception and abortion knowledge, attitudes and practices among adolescents from low and middle-income countries: a systematic review. BMC Health Services Research. 2018;18(1):909–909. doi:10.1186/s12913-018-3722-5

15. Londras FD, School BL. Abortion care guideline Web Annex A. Key international human rights standards on abortion.

16. RODA. Odgovor Vlade RH o slučaju Mirele Čavajde Posebnim procedurama UN za ljudska prava. RODA. Published March 6, 2023. Accessed September 15, 2023. https://www.roda.hr/udruga/projekti/radar/odgovor-vlade-rh-o-slucaju-mirele-cavajde-posebnim-procedurama-un-za-ljudska-prava.html

17. Mellet v. Ireland, 2016; Whelan v. Ireland, 2017 (United Nations Human Rights Committee). Center for Reproductive Rights. Accessed September 15, 2023. https://reproductiverights.org/case/mellet-v-ireland-2016-whelan-v-ireland-2017-united-nations-human-rights-committee/

18. Goldberg J. U.N. Committee Calls for Abortion Law Reform in Chile. Center for Reproductive Rights. Published June 23, 2015. Accessed September 15, 2023. https://reproductiverights.org/u-n-committee-calls-for-abortion-law-reform-in-chile/

19. Austin J, Guy S, Lee-Jones L, McGinn T, Schlecht J. Reproductive Health: A Right for Refugees and Internally Displaced Persons. Reproductive Health Matters. 2008;16(31):10–21.

20. Chynoweth SK. Advancing reproductive health on the humanitarian agenda: the 2012-2014 global review. Conflict and Health. 2015;9(1):I1. doi:10.1186/1752-1505-9-S1-I1

21. Daigle M, Spencer A. Reproductive justice, sexual rights and bodily autonomy in humanitarian action: what a justice lens brings to crisis response. ODI: Think change. Published November 30, 2022. Accessed July 15, 2024. https://odi.org/en/publications/reproductive-justice-sexual-rights-and-bodily-autonomy-in-humanitarian-action-what-a-justice-lens-brings-to-crisis-response/

22. Including refugees and IDPs in national data systems – Forced Migration Review. Accessed July 15, 2024. https://www.fmreview.org/krynskybaal/

23. Casey SE. Evaluations of reproductive health programs in humanitarian settings: a systematic review. Conflict and Health. 2015;9(1):S1. doi:10.1186/1752-1505-9-S1-S1

24. Le Voir R. Leaving no One Behind: Displaced Persons and Sustainable Development Goal Indicators on Sexual and Reproductive Health. Popul Res Policy Rev. 2023;42(5):77. doi:10.1007/s11113-023-09820-z

25. Heidari S, Onyango MA, Chynoweth S. Sexual and reproductive health and rights in humanitarian crises at ICPD25+ and beyond: consolidating gains to ensure access to services for all. Sexual and Reproductive Health Matters. 2019;27(1):343–345. doi:10.1080/26410397.2019.1676513

26. Refugee definition. UNHCR. Published March 1, 2019. Accessed July 14, 2024. https://emergency.unhcr.org/protection/legal-framework/refugee-definition

27. Indicator Metadata Registry Details. Accessed October 17, 2022. https://www.who.int/data/gho/indicator-metadata-registry/imr-details/4622

28. The World’s Abortion Laws. Center for Reproductive Rights. Accessed May 9, 2022. https://reproductiverights.org/maps/worlds-abortion-laws/

29. Abortion Law: Global Comparisons. Council on Foreign Relations. Accessed August 25, 2023. https://www.cfr.org/article/abortion-law-global-comparisons

30. GAPD - The Global Abortion Policies Database - The Global Abortion Policies Database is designed to strengthen global efforts to eliminate unsafe abortion. Accessed September 15, 2023. https://abortion-policies.srhr.org/

31. Wickham H, François R, Henry L, et al. dplyr: A Grammar of Data Manipulation. Published online November 17, 2023. Accessed July 15, 2024. https://cran.r-project.org/web/packages/dplyr/index.html

32. Wickham H, Chang W, Henry L, et al. ggplot2: Create Elegant Data Visualisations Using the Grammar of Graphics. Published online April 23, 2024. Accessed July 15, 2024. https://cran.r-project.org/web/packages/ggplot2/index.html

33. R: A Language and Environment for Statistical Computing. https://www.r-project.org/

34. Zotero | Your personal research assistant. Accessed October 27, 2023. https://www.zotero.org/

35. Tricco AC, Lillie E, Zarin W, et al. PRISMA Extension for Scoping Reviews (PRISMA-ScR): Checklist and Explanation. Ann Intern Med. 2018;169(7):467–473. doi:10.7326/M18-0850

36. Ruzibiza Y. ‘They are a shame to the community…’stigma, school attendance, solitude and resilience among pregnant teenagers and teenage mothers in Mahama refugee camp, Rwanda. Global public health. 2021;16(5):763–774.

37. Williams TP, Chopra V, Chikanya SR. “ It isn’t that we’re prostitutes”: Child protection and sexual exploitation of adolescent girls within and beyond refugee camps in Rwanda. Child Abuse & Neglect. 2018;86:158–166.

38. Ivanova O, Rai M, Mlahagwa W, et al. A cross-sectional mixed-methods study of sexual and reproductive health knowledge, experiences and access to services among refugee adolescent girls in the Nakivale refugee settlement, Uganda. Reprod Health. 2019;16(1):35. doi:10.1186/s12978-019-0698-5

39. Nakisita O, Kirabo-Nagemi C. Examining the maternal health services-seeking behaviour among adolescent urban refugees in Kampala, Uganda. Afr Health Sci. 2023;23(1):394–399. doi:10.4314/ahs.v23i1.41

40. Meyer K, Abimpaye M, Harerimana J de D, Williams C, Gallagher MC. Understanding the Sexual and Reproductive Health Experiences of Refugee and Host Community Adolescents and Youth in Rwanda During COVID-19: Needs, Barriers, and Opportunities. Front Reprod Health. 2022;4:799699. doi:10.3389/frph.2022.799699

41. Asnong C, Fellmeth G, Plugge E, et al. Adolescents’ perceptions and experiences of pregnancy in refugee and migrant communities on the Thailand-Myanmar border: a qualitative study. Reproductive Health. 2018;15:1–13.

42. Napier-Raman S, Hossain SZ, Lee MJ, Mpofu E, Liamputtong P, Dune T. Migrant and refugee youth perspectives on sexual and reproductive health and rights in Australia: a systematic review. Sex Health. 2023;20(1):35–48. doi:10.1071/SH22081

43. Hawkey AJ, Ussher JM, Perz J. Regulation and Resistance: Negotiation of Premarital Sexuality in the Context of Migrant and Refugee Women. J Sex Res. 2018;55(9):1116–1133. doi:10.1080/00224499.2017.1336745

44. Suha M, Murray L, Warr D, et al. Reproductive coercion as a form of family violence against immigrant and refugee women in Australia. PLoS One. 2022;17(11):e0275809. doi:10.1371/journal.pone.0275809

45. Elnakib S, Hussein SA, Hafez S, et al. Drivers and consequences of child marriage in a context of protracted displacement: a qualitative study among Syrian refugees in Egypt. BMC Public Health. 2021;21(1):674. doi:10.1186/s12889-021-10718-8

46. Islam MM, Khan MN, Rahman MM. Factors affecting child marriage and contraceptive use among Rohingya girls in refugee camps. Lancet Reg Health West Pac. 2021;12:100175. doi:10.1016/j.lanwpc.2021.100175

47. Loutet MG, Logie CH, Okumu M, et al. Sexual and reproductive health factors associated with child, early and forced marriage and partnerships among refugee youth in a humanitarian setting in Uganda: Mixed methods findings. Afr J Reprod Health. 2022;26(12s):66–77. doi:10.29063/ajrh2022/v26i12s.8

48. Islam M, Habib SE. “ I don’t want my marriage to end”-A qualitative investigation of the sociocultural factors influencing contraceptive use among married Rohingya women residing in refugee camps in Bangladesh. Published online 2023.

49. Islam MM, Rahman MM, Khan MN. Barriers to male condom use in Rohingya refugee camps in Bangladesh: A qualitative study. Lancet Reg Health Southeast Asia. 2022;2:100008. doi:10.1016/j.lansea.2022.04.004

50. Khan MN, Khanam SJ. Women’s participation in childbearing decision-making and its effects on short-interval births in Rohingya refugee camps of Bangladesh. Lancet Reg Health Southeast Asia. 2023;15:100250. doi:10.1016/j.lansea.2023.100250

51. Hawkey AJ, Ussher JM, Perz J. “If you don’t have a baby, you can’t be in our culture”: migrant and refugee women’s experiences and constructions of fertility and fertility control. Women’s reproductive health. 2018;5(2):75–98.

52. Çöl M, Bilgili Aykut N, Usturalı Mut AN, et al. Sexual and reproductive health of Syrian refugee women in Turkey: a scoping review within the framework of the MISP objectives. Reprod Health. 2020;17(1):99. doi:10.1186/s12978-020-00948-1

53. Alan Dikmen H, Cankaya S, Dereli Yilmaz S. The attitudes of refugee women in Turkey towards family planning. Public Health Nursing. 2019;36(1):45–52.

54. Abul Kalam Azad Md, Zakaria M, Nachrin T, Chandra Das M, Cheng F, Xu J. Family planning knowledge, attitude and practice among Rohingya women living in refugee camps in Bangladesh: a cross-sectional study. Reproductive Health. 2022;19(1):105. doi:10.1186/s12978-022-01410-0

55. Hossain MA, Hossain MB. Understanding fertility behavior of the Forcibly Displaced Myanmar Nationals in Bangladesh: A qualitative study. PLoS One. 2023;18(5):e0285675. doi:10.1371/journal.pone.0285675

56. Royer PA, Olson LM, Jackson B, et al. “In Africa, There Was No Family Planning. Every Year You Just Give Birth”: Family Planning Knowledge, Attitudes, and Practices Among Somali and Congolese Refugee Women After Resettlement to the United States. Qual Health Res. 2020;30(3):391–408. doi:10.1177/1049732319861381

57. Gee S, Vargas J, Foster AM. “The more children you have, the more praise you get from the community”: exploring the role of sociocultural context and perceptions of care on maternal and newborn health among Somali refugees in UNHCR supported camps in Kenya. Confl Health. 2019;13:11. doi:10.1186/s13031-019-0195-z

58. Zhang Y, McCoy EE, Scego R, Phillips W, Godfrey E. A Qualitative Exploration of Somali Refugee Women’s Experiences with Family Planning in the U.S. J Immigrant Minority Health. 2020;22(1):66–73. doi:10.1007/s10903-019-00887-5

59. Agbemenu K, Volpe EM, Dyer E. Reproductive health decision-making among US- dwelling Somali Bantu refugee women: A qualitative study. Journal of Clinical Nursing. 2018;27(17-18):3355–3362.

60. Singh NS, Prabhakar P, Ssali A, et al. “They will say you want to make their home die”: A mixed methods study to assess modern family planning use in partnered South Sudanese refugee and host populations in Northern Uganda. PLOS Glob Public Health. 2022;2(6):e0000348. doi:10.1371/journal.pgph.0000348

61. Mourtada R, Melnikas AJ. Crisis upon crisis: a qualitative study exploring the joint effect of the political, economic, and pandemic challenges in Lebanon on Syrian refugee women’s fertility preferences and behaviour. Confl Health. 2022;16(1):35. doi:10.1186/s13031-022-00468-8

62. Odwe G, Undie CC, Obare F. Attitudes towards help-seeking for sexual and gender-based violence in humanitarian settings: the case of Rwamwanja refugee settlement scheme in Uganda. BMC international health and human rights. 2018;18:1–12.

63. Coşkun AM, Özerdoğan N, Karakaya E, Yakıt E. Fertility characteristics and related factors impacting on Syrian refugee women living in Istanbul. Afr Health Sci. 2020;20(2):682–689. doi:10.4314/ahs.v20i2.19

64. Freedman J, Crankshaw TL, Mutambara VM. Sexual and reproductive health of asylum seeking and refugee women in South Africa: understanding the determinants of vulnerability. Sex Reprod Health Matters. 2020;28(1):1758440. doi:10.1080/26410397.2020.1758440

65. Donnelly A, Haintz GL, McKenzie H, Graham M. Influences on reproductive decision-making among forcibly displaced women resettling in high-income countries: a scoping review and thematic analysis. International Journal for Equity in Health. 2023;22(1):179. doi:10.1186/s12939-023-01993-5

66. Chalmiers MA, Karaki F, Muriki M, Mody SK, Chen A, Thiel de Bocanegra H. Refugee women’s experiences with contraceptive care after resettlement in high-income countries: A critical interpretive synthesis. Contraception. 2022;108:7–18. doi:10.1016/j.contraception.2021.11.004

67. AlArab N, Nabulsi D, El Arnaout N, et al. Reproductive health of Syrian refugee women in Lebanon: a descriptive analysis of the Sijilli electronic health records database. BMC Womens Health. 2023;23(1):81. doi:10.1186/s12905-023-02231-4

68. Kingori C, Ice GH, Hassan Q, Elmi A, Perko E. ‘If I went to my mom with that information, I’m dead’: sexual health knowledge barriers among immigrant and refugee Somali young adults in Ohio. Ethnicity & health. 2018;23(3):339–352.

69. Persson M, Larsson EC, Islam NP, Gemzell-Danielsson K, Klingberg-Allvin M. A qualitative study on health care providers’ experiences of providing comprehensive abortion care in Cox’s Bazar, Bangladesh. Confl Health. 2021;15(1):6. doi:10.1186/s13031-021-00338-9

70. Tanabe M, Greer A, Leigh J, et al. An exploration of gender-based violence in eastern Myanmar in the context of political transition: findings from a qualitative sexual and reproductive health assessment. Sex Reprod Health Matters. 2019;27(2):1665161. doi:10.1080/26410397.2019.1665161

71. Soin KS, Beldowski K, Bates E, et al. Attitudes Towards Family Planning among Bhutanese, Burmese, and Iraqi Refugee Women: A Qualitative Study. Hawaii J Health Soc Welf. 2020;79(6 Suppl 2):70–77.

72. Maheen H, Chalmers K, Khaw S, McMichael C. Sexual and reproductive health service utilisation of adolescents and young people from migrant and refugee backgrounds in high-income settings: a qualitative evidence synthesis (QES). Sex Health. 2021;18(4):283–293. doi:10.1071/SH20112

73. Sawadogo PM, Sia D, Onadja Y, et al. Barriers and facilitators of access to sexual and reproductive health services among migrant, internally displaced, asylum seeking and refugee women: A scoping review. PLoS One. 2023;18(9):e0291486. doi:10.1371/journal.pone.0291486

74. Besera G, Vu M, Dogbe A, et al. “My culture doesn’t 100% like these kinds of services, but you decide what to do”: Female refugees’ experiences with sexual and reproductive healthcare in the Southeastern U.S. PEC Innov. 2023;2:100172. doi:10.1016/j.pecinn.2023.100172

75. Mourtada R, Melnikas AJ. Syrian refugee women’s access to family planning services and modern contraception during overlapping crises in Bekaa, Lebanon. BMC Womens Health. 2023;23(1):475. doi:10.1186/s12905-023-02613-8

76. Márquez-Lameda RD. Predisposing and enabling factors associated with Venezuelan migrant and refugee women’s access to sexual and reproductive health care services and contraceptive usage in Peru. J Migr Health. 2022;5:100107. doi:10.1016/j.jmh.2022.100107

77. Fouad FM, Hashoush M, Diab JL, et al. Perceived facilitators and barriers to the provision of sexual and reproductive health services in response to the Syrian refugee crisis in Lebanon. Womens Health (Lond). 2023;19:17455057231171486. doi:10.1177/17455057231171486

78. Munyaneza Y, Mhlongo EM. Challenges of women refugees in utilising reproductive health services in public health institutions in Durban, KwaZulu-Natal, South Africa. Health SA. 2019;24:1030. doi:10.4102/hsag.v24i0.1030

79. Gebreyesus T, Gottlieb N, Sultan Z, et al. Barriers to contraceptive careseeking: the experience of Eritrean asylum-seeking women in Israel. Ethn Health. 2020;25(2):255–272. doi:10.1080/13557858.2017.1418299

80. Nara R, Banura A, Foster AM. Assessing the availability and accessibility of emergency contraceptive pills in Uganda: A multi-methods study with Congolese refugees. Contraception. 2020;101(2):112–116. doi:10.1016/j.contraception.2019.09.008

81. Muuo S, Muthuri SK, Mutua MK, et al. Barriers and facilitators to care-seeking among survivors of gender-based violence in the Dadaab refugee complex. Sexual and reproductive health matters. 2020;28(1):1722404.

82. Banke-Thomas A, Agbemenu K, Johnson-Agbakwu C. Factors Associated with Access to Maternal and Reproductive Health Care among Somali Refugee Women Resettled in Ohio, United States: A Cross-Sectional Survey. J Immigr Minor Health. 2019;21(5):946–953. doi:10.1007/s10903-018-0824-4

83. Arnott G, Otema C, Obalim G, Odallo B, Nakubulwa T, Okello SBT. Human rights-based accountability for sexual and reproductive health and rights in humanitarian settings: Findings from a pilot study in northern Uganda. PLOS Glob Public Health. 2022;2(8):e0000836. doi:10.1371/journal.pgph.0000836

84. Korri R, Froeschl G, Ivanova O. A Cross-Sectional Quantitative Study on Sexual and Reproductive Health Knowledge and Access to Services of Arab and Kurdish Syrian Refugee Young Women Living in an Urban Setting in Lebanon. Int J Environ Res Public Health. 2021;18(18):9586. doi:10.3390/ijerph18189586

85. Amiri M, El-Mowafi IM, Chahien T, Yousef H, Kobeissi LH. An overview of the sexual and reproductive health status and service delivery among Syrian refugees in Jordan, nine years since the crisis: a systematic literature review. Reprod Health. 2020;17(1):166. doi:10.1186/s12978-020-01005-7

86. Nara R, Banura A, Foster AM. Exploring Congolese refugees’ experiences with abortion care in Uganda: a multi-methods qualitative study. Sex Reprod Health Matters. 2019;27(1):1681091. doi:10.1080/26410397.2019.1681091

87. Uganda’s Abortion Provisions. Center for Reproductive Rights. Accessed March 11, 2023. https://reproductiverights.org/maps/provision/ugandas-abortion-provisions/

88. Schulte-Hillen C, Staderini N, Saint-Sauveur JF. Why Médecins Sans Frontières (MSF) provides safe abortion care and what that involves. Confl Health. 2016;10:19. doi:10.1186/s13031-016-0086-5

89. Kumar M, Schulte-Hillen C, De Plecker E, et al. Catalyst for change: Lessons learned from overcoming barriers to providing safe abortion care in Médecins Sans Frontières projects. Perspect Sex Reprod Health. Published online October 23, 2022. doi:10.1363/psrh.12209

90. McGinn T, Casey SE. Why don’t humanitarian organizations provide safe abortion services? Confl Health. 2016;10(1):8. doi:10.1186/s13031-016-0075-8

91. Uddin N. The State, Vulnerability, and Transborder Movements: The Rohingya People in Myanmar and Bangladesh. In: Uddin N, Chowdhory N, eds. Deterritorialised Identity and Transborder Movement in South Asia. Springer; 2019:73–90. doi:10.1007/978-981-13-2778-0_5

92. Ekmekci PE. Syrian Refugees, Health and Migration Legislation in Turkey. J Immigr Minor Health. 2017;19(6):1434–1441. doi:10.1007/s10903-016-0405-3

93. Yaya S, Odusina EK, Bishwajit G. Prevalence of child marriage and its impact on fertility outcomes in 34 sub-Saharan African countries. BMC international health and human rights. 2019;19(1):1–11.

94. Tinago CB, Annang Ingram L, Frongillo EA, Simmons D, Blake CE, Engelsmann B. Understanding the Social Environmental Influences on Pregnancy and Planning for Pregnancy for Young Women in Harare, Zimbabwe. Matern Child Health J. 2019;23(12):1679–1685. doi:10.1007/s10995-019-02814-4

95. Pasquier E, Owolabi OO, Fetters T, et al. High severity of abortion complications in fragile and conflict-affected settings: a cross-sectional study in two referral hospitals in sub-Saharan Africa (AMoCo study). BMC Pregnancy and Childbirth. 2023;23(1):143. doi:10.1186/s12884-023-05427-6

96. State of World Population 1999. United Nations Population Fund. Accessed November 4, 2023. https://www.unfpa.org/publications/state-world-population-1999

